# USe of Diagnostic sUbtraction angiography in The isCHemic stroke setting (US DUTCH study)

**DOI:** 10.1101/2023.10.10.23296855

**Authors:** Maxim J.H.L. Mulder, Diederik W.J. Dippel, James Burke

**Affiliations:** Erasmus University Medical Center, Rotterdam, The Netherlands; The Ohio State University Wexner Medical Center, Columbus, Ohio, United States

## Abstract

**Introduction:** There are no recommendations for DSA in the ischemic stroke work-up according to current guidelines. We studied the rate of DSA in ischemic stroke, the recent time-trend (given the recent rapid increase in mechanical thrombectomy), hospital variation and associated factors.

**Methods:** This is a retrospective cross-sectional study among Medicare fee-for-service beneficiaries with ischemic stroke admitted between 2016 and 2020 in the United States. ICD-10 codes were used to determine ischemic stroke diagnosis and procedure codes for thrombectomy and DSA. Hospital trends and factors associated with DSA performance were analyzed in hospitals with DSA capacity.

**Results:** 7.373 (0.7%) of the 1,085,644 ischemic stroke patients, had a DSA for diagnostic purposes. In the patients that were admitted to a hospital with DSA facility, the following factors showed the strongest association with DSA: younger age (aOR=0.81 [95% confidence interval (CI):0.81-0.83]), thrombectomy rate in that hospital (aOR=2549 [95%CI:610-10663]), transfer (aOR=1.41[95%CI:1.34-1.50]) and carotid disease (aOR=5.8 [95%CI:5.6-6.1]). There was large variation in the hospital DSA rate, varying from 0.07% to 11.1%. Of the variance of DSA rates, 15% was attributed to the residual effect hospital propensity to perform DSA. The top decile of hospitals with the highest DSA rate, performed DSA’s in >2.3% of patients, compared to the 0.6% median. There was no change in DSA rates over time.

**Conclusion:** DSA is used infrequently in acute ischemic stroke patients and did not change between 2016 to 2020. Hospital variation in DSA use was however large, and not solely explained by patient and facility factors.

## Introduction

Digital subtraction angiography (DSA) is the oldest technique for cerebral vessel imaging. DSA is the optimal technique for aneurysm screening after a subarachnoid hemorrhage and is often used to screen for certain other intracranial vascular anomalies. DSA was also used for carotid stenosis grading in the large carotid surgery trials during the 1980s and 1990s, and therefore deemed as the gold standard for identifying and characterizing vascular disease in the ischemic stroke work-up. DSA has a substantial downside, though, since the invasiveness of this imaging technique is associated with complications including: mortality, dissections, groin hematoma and stroke. Published stroke rates caused by DSA varies from 0% to 3.8% in the largest retrospective study (of more than 400,000 patients in the United States).^1, 2^ Stroke risk of DSA varies largely with age and indication, were the highest stroke risk is found in old age and in ischemic stroke patients.^1, 3^ Since non-invasive vascular imaging bears no stroke complication risk and is shown to be cost-effective, these techniques have largely replaced DSA in ischemic stroke work-up.^4, 5^ Ultrasound, CTA or MRA is currently recommended as routine work-up in ischemic stroke patients, according to the American and European guidelines. Neither guideline states in what scenarios DSA should be considered or omitted.^6, 7^

While the utility of DSA remains unclear, the recent introduction of mechanical thrombectomy (MT) for patients with large vessel occlusion has resulted in rapidly growing expertise in both invasive and non-invasive cerebral vessel imaging for ischemic stroke. Many ischemic stroke patients now receive non-invasive imaging of the cerebral vasculature (MR-/CT-angiography) at the emergency department, to screen for large vessel occlusion. The recent and rapid increase in MT may have led to an increase in DSA use, either because of increased DSA availability or because of shifting provider beliefs about the utility of DSAs in the context of more commonly acquiring this information in the context of MT. Conversely, since non-invasive vessel imaging is become more widely available, this could conceivably result in lower DSA rates over time.

The primary aims of this study is to describe the recent rate and temporal trend of DSA for diagnostic work-up of ischemic stroke patients. Secondarily, we explore patient and facility factors associated with DSA and characterize hospital variation in DSA performance.

## Methods

### Study Overview and Key Measures

This is a retrospective cross-sectional study characterizing: 1. Overall DSA rates 2. Temporal trends in DSA rates 3. Patient and facility factors associated with DSA and 4. Facility-level variation in DSA among Medicare fee-for-service beneficiaries. We used Medicare inpatient files from 2016–2020 to identify all patients with a primary diagnosis of ischemic stroke admitted to an acute care hospital using ICD-10 codes, defined as an I63 code in the primary diagnosis with the exception of I363 ‘Cerebral infarction due to cerebral venous thrombosis, nonpyogenic’.^8^ See supplemental table 4 for the definition of all analyzed variables. Facility information was derived from inpatient files (e.g. stroke volume by calculated by summing the number of inpatient stroke hospitalizations in a given year) or by using Medicare hospital provider cost report hospital data (2019).

MT and carotid endarterectomy (CEA) procedures were identified using ICD-10 procedure codes in the inpatient file (supplemental table 4). For MT we used all 03CG3-03CQ3 codes and for CEA all 03CG0-03CQ0 codes.^9^ For MT, MS-DRG codes 0.23 or 0.24 were also used (when not having a simultaneous ICD-10-PCS code for craniotomy, craniectomy or ventriculostomy), since the sensitivity of ICD procedure codes in isolation for MT is suboptimal, 86.4%.^8^ Many MT and carotid endovascular procedures were also coded as having DSA on the same day. Since our aim was to analyze DSA procedures that were done for diagnostic purposes only, we limited the DSA definition to capture cases most likely to represent diagnostic DSA — DSAs that were performed at least two days after the stroke admission date, and not on the same date as MT or a carotid endovascular procedure (supplemental table 4). The reason for the 2-day delay was to exclude MT procedures (which may occur on the day of admission or the subsequent day) and repeat procedures that may be related to the MT itself.

### Statistical analysis

We sought to characterize 4 key concepts: 1. The proportion of ischemic stroke patients in whom a DSA was performed for diagnostic purposes. This proportion was separately calculated for all ischemic stroke patients in our sample and for all ischemic stroke patients that were seen in a hospital with the capacity to perform a DSA. Descriptive statistics were used to compare patients that had a DSA performed and those who did not. Chi-square test was used for categorical variables. For continuous variables we used the t-test when normally distributed and Wilcoxon-Mann-Whitney test when this was not the case; 2. Time trends in the use of DSA per year were estimated as absolute percentages of all ischemic stroke patients and the population that was seen in hospitals with DSA facilities. Also, year was used in the regression analysis to determine a possible association with DSA (see the following point); 3. Patient and hospital factors associated with DSA were analyzed amongst subjects treated in a DSA capable hospital (DSA treated hospital was defined as) using multi-level logistic regression for the dependent variable, DSA and including the independent variables listed below as well as random facility level intercept. We explored the association of the following patient and hospital factors with DSA use:

-age
- sex
- race
- comorbidities: hypertension, hyperlipidemia, diabetes, smoking, atrial fibrillation/flutter, myocardial infarct, heart failure, kidney failure, peripheral artery disease, dementia, COPD, COPD, cancer, dementia
- carotid stenosis/occlusion
- specific stroke diagnosis (dissection, Moyamoya disease, cerebral arteritis, reversible cerebrovascular vasoconstriction syndrome or other cerebrovascular vasospasm and vasoconstriction)
- performance of IVT in that patient
- performance of IAT in that patient
- year
- hospital transfer
- percentage of IVTs in all ischemic stroke patients in that hospital
- percentage of IATs in all ischemic stroke patients in that hospital
- profit making hospital
- urban hospital
- teaching hospital
- number of beds in that hospital
- number of ischemic stroke in that hospital per year
- hospital center

4. Facility level variation in the use of DSA was characterized using the intraclass correlation coefficient from the model in #3 after adjustment for all other patient and facility factors. Facility effects were characterized with caterpillar plots of the predicted probability of performing DSA within a given facility. To gain more insight in hospital variation we describe baseline variables between hospitals with the highest decile of DSA rate, and the other DSA hospitals.

Since this is a cross-sectional study using data of routinely collected health data (Medicare), we adhered to the STROBE guidelines with RECORD extension.^10^ All analysis were completed in Stata.(Release 15.1. 2017 StataCorp LLC).

### Sensitivity Analyses

To assess the robustness of our primary analyses, we performed several sensitivity analyses. First, we assessed whether DSA coding may be omitted in the inpatient file, based on ICD-10 coding entered by hospital coders, compared to DSAs coded in the Carrier file, based on CPT coding (36211-36228) at the provider level. We made this comparison in 2016 year, since that year the inpatient DSA rate was lowest and therefore most likely to yield discrepancies. We used the claim carrier file to filter for stroke diagnosis, and then linked these to the procedure codes. Furthermore we explored the associations of the above mentioned patient and hospital characteristics with IVT and MT and we performed this analysis on the patients that were coded as having a DSA within 2 days after stroke.

## Results

### DSA rate

Between 2016 and 2020, a total of 1,085,644 Medicare fee-for-service beneficiaries were hospitalized with acute ischemic stroke as the primary diagnosis. Figure 1 shows the inclusion/exclusion flowchart defining the population of ischemic stroke patients that likely had a DSA for diagnostic purposes. In total, 52,519 patients (4.8%) had a DSA ICD-10 procedure code. Of them 28,265 (53.8%) were coded as having a MT and 3,544 (6.8%) a carotid endovascular procedure (stent and/or percutaneous transluminal angioplasty) on the same date. In our cohort 5% of MT procedures were only coded using MS-DRG. Of the remaining 20,710 patients with a DSA coded, 12,873 had the DSA (62.2%) performed on the day of stroke or the day after and 464 patients (2.2%) before the stroke date. So the remaining 7,373 patients (0.7%), were defined as having a DSA for diagnostic purposes. In most patients (92.7%) the DSA was performed in the week after stroke (Supplemental figure 1). Of all 4,382 hospitals, 49.1% performed IVT, 20.6% performed MT and 20.8% performed DSA. In the hospitals with DSA facilities, DSA rate was 1.1%.

**Figure 1.**
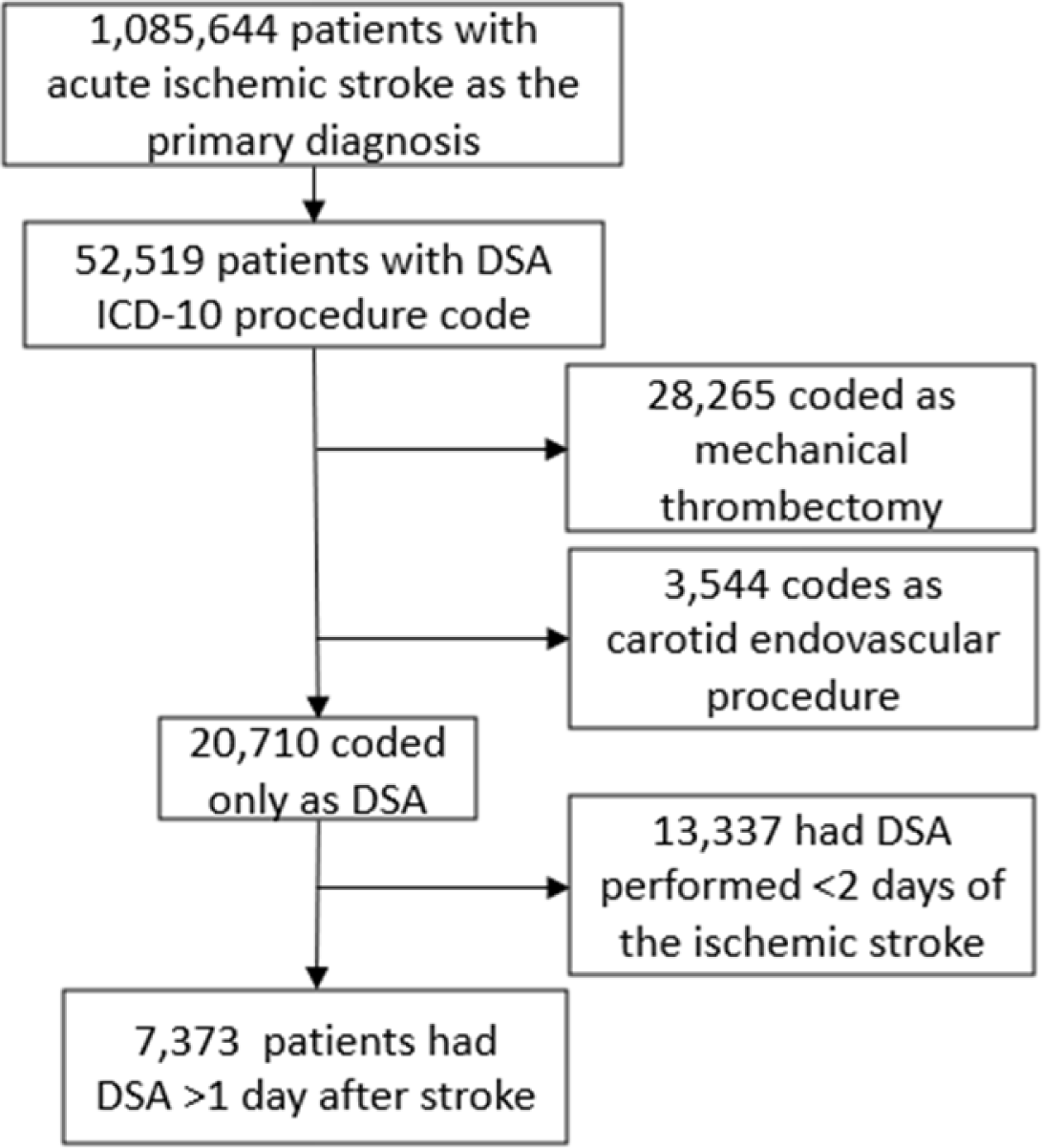
Flowchart of ischemic stroke patients coded as DSA and used for analyses.

### Time trend in DSA

During the study period from 2016 to 2020 the rate of DSA in ischemic stroke patients was stable, while the MT rate more than doubled (figure 2). This persisted in multi variable analyses accounting for time independently of all other patient and facility factors studied.

**Figure 2.**
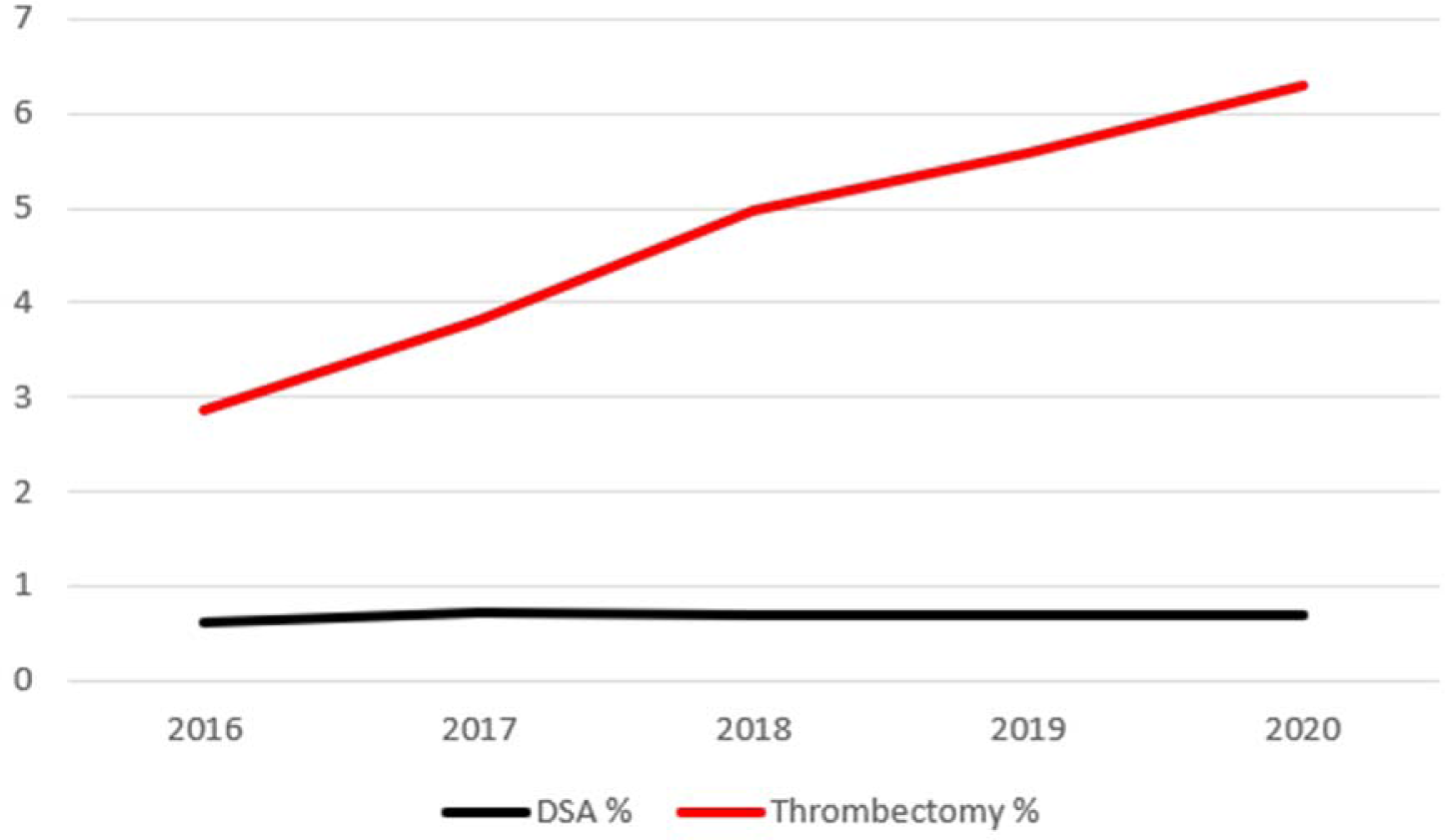
Time trend of all ischemic stroke patients that underwent DSA or thrombectomy per year.

### Patient and Facility Factors Associated with DSA

There were many differences between patients that had a DSA and those who did not, in hospitals with DSA facilities (table 1). This was largely comparable with patients that were treated in all hospitals, with and without DSA facilities (Supplemental table1). A number of patient factors were independently associated with DSA (table 2): younger age, male gender, atherosclerotic comorbidities, renal failure, peripheral vascular disease and specific stroke diagnosis (dissection, Moyamoya disease, cerebral arteritis, reversible cerebrovascular vasoconstriction syndrome or other cerebrovascular vasospasm and vasoconstriction). The factor with the largest difference between the DSA and non-DSA groups was suspected carotid disease (stenosis or occlusion), 61% in the DSA group vs. 20% in the non-DSA group (p<0.01). Factors associated with not performing DSA were: atrial fibrillation, cancer and dementia. The following facility factors were associated with a higher DSA rate in after adjustments: transferred patients, higher MT rate in that facility and profit hospital (Table 2). The factors with the strongest associations were; younger age (aOR=0.81 [95% confidence interval (CI):0.81-0.83]), thrombectomy rate in that hospital (aOR=2549 [95%CI:610-10663]), transfer (aOR=1.41[95%CI:1.34-1.50]) and carotid disease (aOR=5.8 [95%CI:5.6-6.1]).(see figure 3 & 4).

**Table 1.** Demographics, comorbidities and hospital factors by patients with and without a DSA performed in hospitals that ware capable of performing DSA.

| *Demographics* | **DSA performed**<br>n=7.373 | **No DSA performed**<br>n=672.648 | **p-value** |
| --- | --- | --- | --- |
| Age <65 | 1.181 (16.0%) | 66.024 (9.8%) | <0.01 |
| 65-69 | 1.739 (23.6%) | 109.703 (16.4%) |  |
| 70-74 | 1.561 (21.2%) | 110.608 (16.4%) |  |
| 75-79 | 1.277 (17.3%) | 109.990 (16.4%) |  |
| 80-84 | 965 (13.1%) | 105.892 (15.7%) |  |
| >84 | 652 (8.8%) | 170.431 (25.3%) |  |
| Male | 4.133 (56.0%) | 316.580 (47.1%) | <0.01 |
| Race: White | 5.773 (78.3%) | 531.049 (79.0%) | <0.01 |
| Black | 1.039 (14.1%) | 95.432 (14.2%) |  |
| Other | 130 (1.8%) | 10.734 (1.6%) |  |
| Asian | 117 (1.6%) | 12.725 (1.9%) |  |
| Hispanic | 159 (2.2%) | 12.217 (1.8%) |  |
| Native American | 62 (0.8%) | 3.699 (0.6%) |  |
| *Comorbidities* |  |  |  |
| Hypertension | 4.360 (59.1%) | 385.308 (57.3%) | <0.01 |
| Hyperlipidemia | 5.206 (70.6%) | 430.259 (64.0%) | <0.01 |
| Diabetes | 3.269 (44.3%) | 258.278 (38.4%) | <0.01 |
| Smoking | 1.582 (21.5%) | 95.317 (14.2%) | <0.01 |
| Atrial fibrillation/flutter | 1.608 (21.8%) | 230.792 (34.3%) | <0.01 |
| Myocardial infarction | 247 (3.4%) | 23.749 (3.5%) | 0.40 |
| Heart failure | 1.291 (17.5%) | 133.001 (19.8%) | <0.01 |
| Renal failure | 2.212 (30.0%) | 196.534 (29.2%) | 0.15 |
| Peripheral vascular disease | 548 (7.4%) | 34.871 (5.2%) | <0.01 |
| COPD | 1.207 (16.4%) | 92.750 (13.8%) | <0.01 |
| Cancer | 544 (7.4%) | 103.332 (15.4%) | <0.01 |
| Dementia | 322 (4.4%) | 37.896 (5.6%) | <0.01 |
| Carotid disease | 4.523 (61.3%) | 135.324 (20.1%) | <0.01 |
| Specific stroke diagnosis | 181 (2.5%) | 1.396 (0.2%) | <0.01 |
| IV thrombolysis | 906 (12.3%) | 100.033 (14.9%) | <0.01 |
| Thrombectomy | 429 (5.8%) | 48.598 (7.2%) | <0.01 |
| *Hospital factors* |  |  |  |
| Transferred patients | 2.454 (33.3%) | 140.608 (20.9%) | <0.01 |
| IVT % per hospital (mean) | 16.1 | 14.8 | <0.01 |
| Thrombectomy % per hospital (mean) | 9.4 | 7.2 | <0.01 |
| Urban hospital | 5.635 (76.4%) | 517.943 (77%) | 0.23 |
| Profit hospital | 934 (12.7%) | 86.009 (12.8%) | 0.76 |
| Teaching hospital | 4.639 (62.9%) | 352.436 (52.4%) | <0.01 |
| Number of beds (mean) | 551 (±324) | 522 | <0.01 |
| Number of ischemic strokes per year (mean) | 252 (±167) | 230 | <0.01 |

**Table 2.** Associations with performance of DSA, adjusted for multiple variables.

|  | **aOR (95% CI)** | **p-value** |
| --- | --- | --- |
| Age | 0.81 (0.81-0.83) | <0.01 |
| Gender (female) | 0.91 (0.87-0.95) | <0.01 |
| Race: White | 1.03 (0.82-1.24) | 0.96 |
| Black | 0.93 (0.75-1.16) | 0.54 |
| Other | 1.12 (0.85-1.47) | 0.43 |
| Asian | 0.92 (0.69-1.21) | 0.54 |
| Hispanic | 0.97 (0.74-1.27) | 0.85 |
| Native American | 1.04 (0.74-1.46) | 0.82 |
| Hypertension | 1.03 (0.97-1.10) | 0.34 |
| Hyperlipidemia | 1.19 (1.13-1.25) | <0.01 |
| Diabetes | 1.05 (1.00-1.11) | 0.05 |
| Smoking | 0.96 (0.90-1.02) | 0.18 |
| Atrial fibrillation/ flutter | 0.72 (0.68-0.76) | <0.01 |
| Myocardial infarction | 0.92 (0.80-1.05) | 0.21 |
| Heart failure | 0.98 (0.91-1.05) | 0.50 |
| Renal failure | 1.11 (1.04-1.18) | <0.01 |
| Peripheral vascular disease | 1.15 (1.05-1.26) | <0.01 |
| COPD | 1.05 (0.98-1.12) | 0.14 |
| Cancer | 0.81 (0.73-0.92) | <0.01 |
| Dementia | 0.68 (0.92-0.74) | <0.01 |
| Carotid disease | 5.84 (5.56-6.13) | <0.01 |
| Specific stroke diagnosis | 8.53 (7.15-10.2) | <0.01 |
| IV thrombolysis | 0.77 (0.71-0.83) | <0.01 |
| Thrombectomy | 0.49 (0.44-0.54) | <0.01 |
| Year of stroke | 1.01 (0.99-1.03) | 0.17 |
| Transfer | 1.41 (1.34-1.50) | <0.01 |
| IVT % (hospital) | 0.08 (0.02-0.30) | <0.01 |
| Thrombectomy % (hospital) | 2549.56 (609.62-10662.72) | <0.01 |
| Urban hospital | 1.03 (0.89-1.20) | 0.66 |
| Profit hospital | 1.29 (1.10-1.52) | <0.01 |
| Teaching hospital | 1.15 (0.99-1.33) | 0.07 |
| Number of beds | 1.00 (1.00-1.00) | 0.07 |
| Number of ischemic strokes per year | 1.00 (1.00-1.00) | 0.58 |

**Figure 3.**
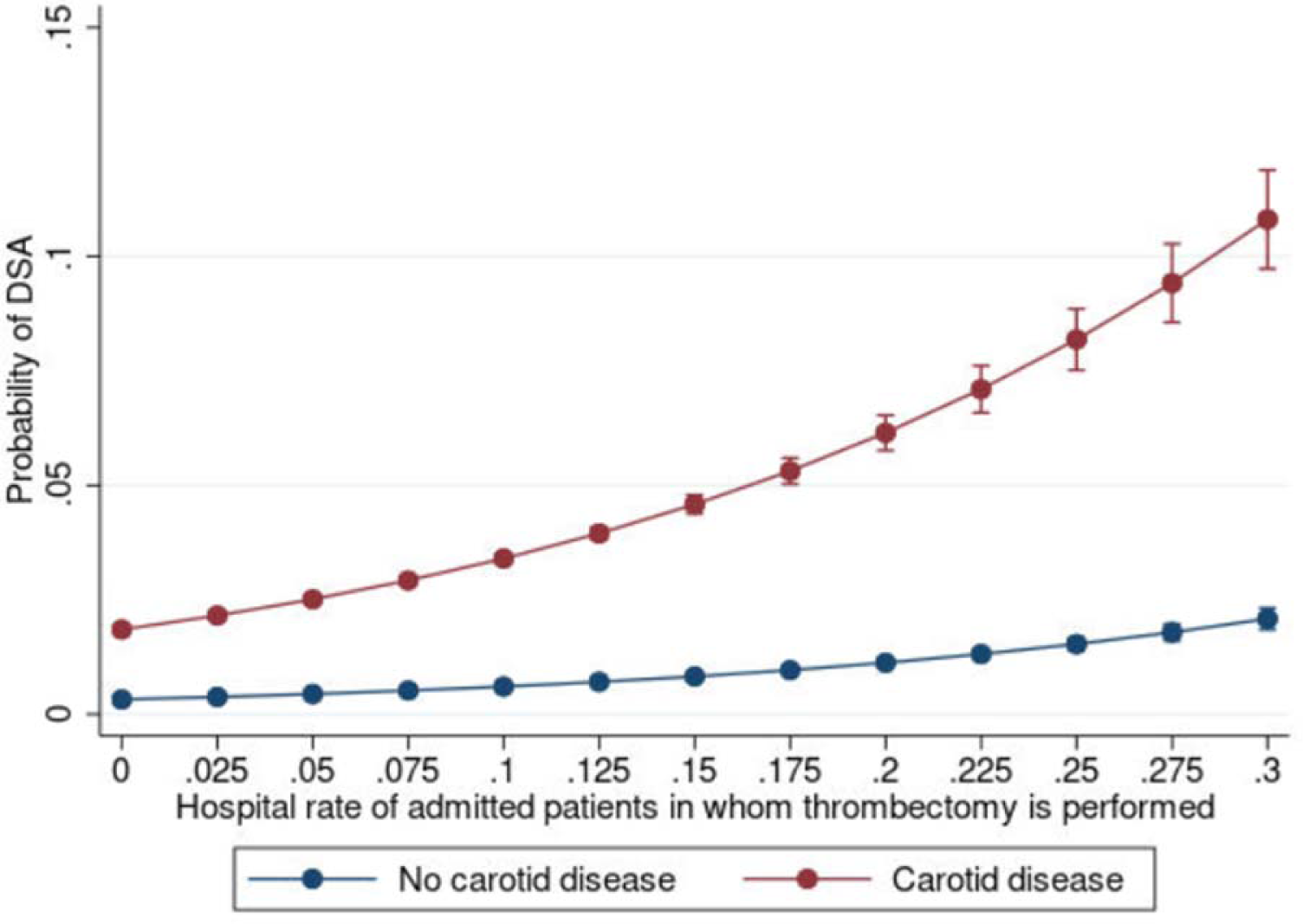
Probability of DSA with increasing % of patients who get thrombectomy in that hospital (x-axis), for patients with carotid stenosis/occlusions (red line) and patients without carotid stenosis/occlusion (blue line).

**Figure 4.**
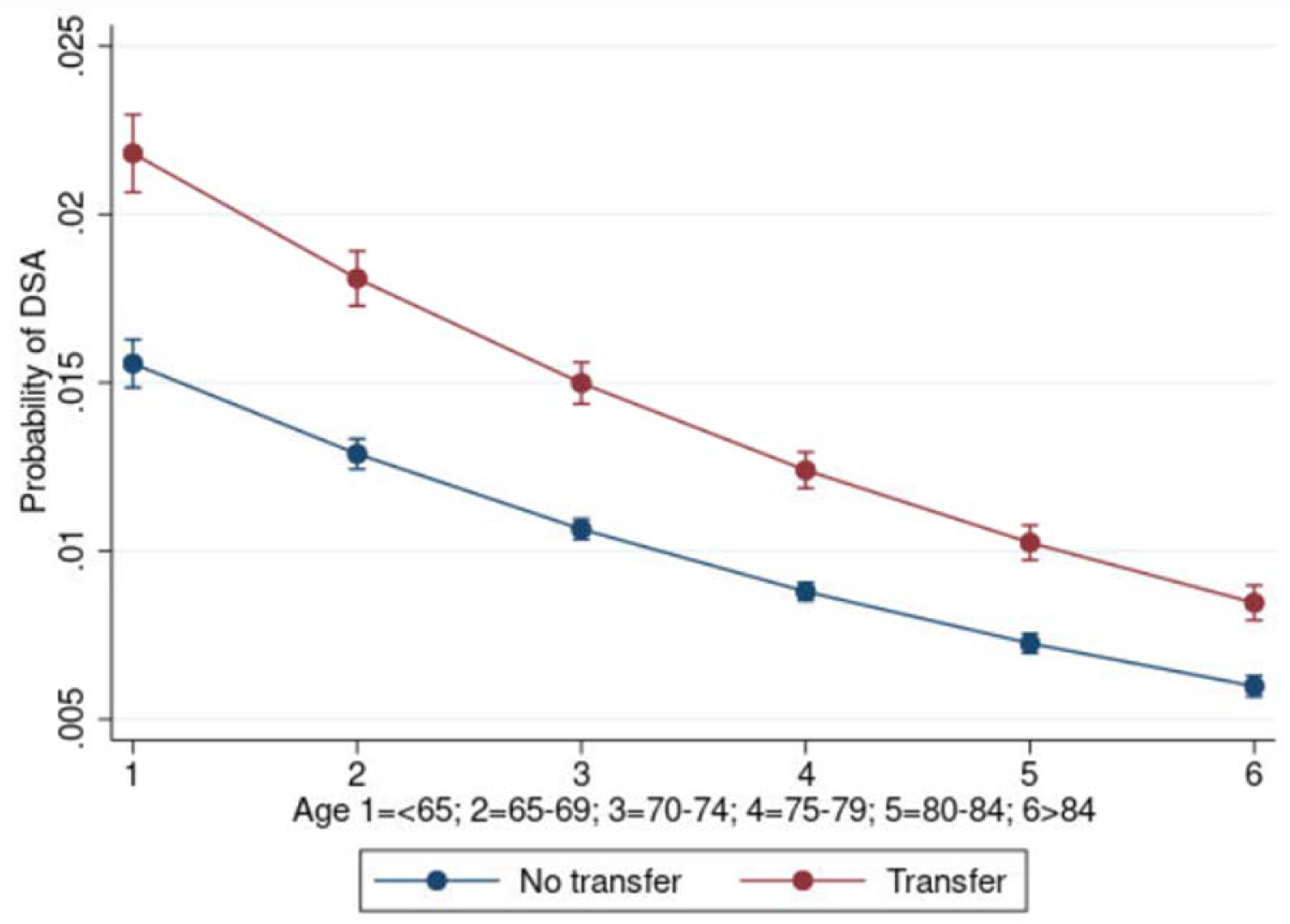
Probability of DSA with age (x-axis), for patients who were transferred from another hospital (red line) and patients that were not transferred (blue line).

Supplemental table 2 shows outcome related factors of patients that underwent DSA and who did not. There was a substantial differences in carotid procedures in the DSA group (16% vs. 3%). Furthermore hospital stay was longer in DSA patients, but less patients died during hospital admission (Supplemental table 2).

### Facility Variation in the use of DSA

There were 910 hospitals that performed DSA (20.8%). These higher volume hospitals cared for 682,188 ischemic stroke patients, representing 62.8% of the entire sample. In these hospitals the DSA rate was 1.1% (7,373 patients). The DSA rate per hospital ranged from 0.07% to 11.1%. Most hospitals performed DSA in <1% (see table 3). Figure 5 shows the distribution of hospitals and their DSA rate. This shows that a small proportion of hospitals perform DSA in a higher than average proportion of patients. The top decile of DSA performing hospitals (91 hospitals), performed DSA’s in >2.3% of patients, compared to the 0.6% median. Facilities in the top decile had on average more ischemic stroke Medicare fee for service patients per year (174 patients). Patient demographics, comorbidities and hospital factors show that patients in the top decile of DSA rate, were largely similar (Supplemental table 3). Of the patient demographics there were some crude differences were highest percentile DSA rate hospitals patients had higher rate of carotid disease (23% vs. 20%), and more often underwent IVT and/or MT. Hospital factors showed more transfers, higher MT rates and higher stroke volume.

**Table 3.** Number of DSA hospitals per % of hospital DSA rate. Total number of DSA hospitals is 910.

| **DSA rate** | **Number of hospitals (%)** |
| --- | --- |
| >0.5% | 553 (61%) |
| >1% | 324 (36%) |
| >2% | 119 (13%) |
| >3% | 58 (6%) |
| >4% | 28 (3%) |

**Figure 5.**
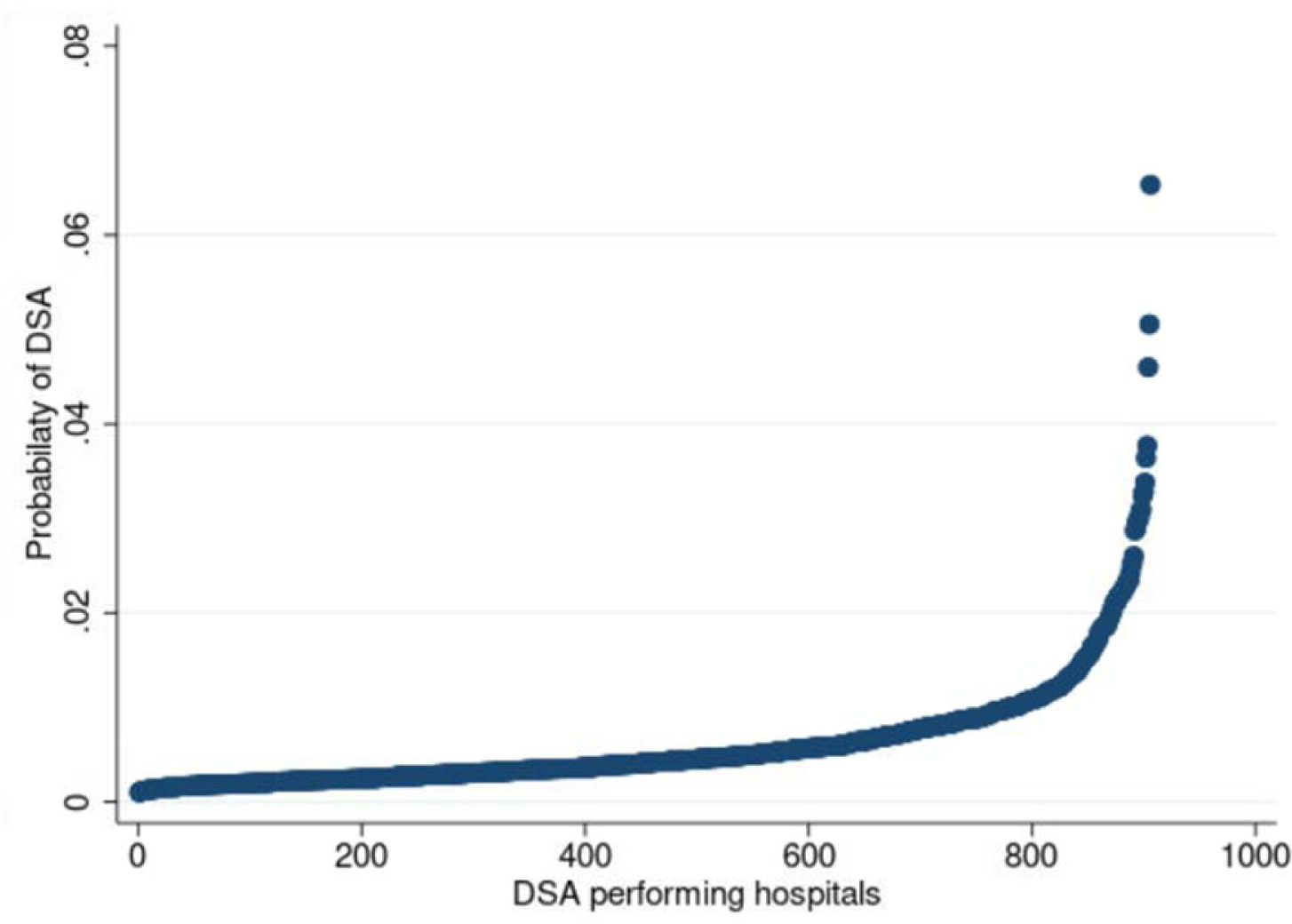
DSA % in all ischemic stroke patients per hospital, from lowest to highest DSA performing hospital (plotted for DSA performing hospitals).

The ICC of hospital on DSA decreased from 0.18 (95% CI: 0.16-0.20) after adjustment with patient factors only, to 0.15 (95% CI: 0.13-0.16) when hospital factors were added. This means that independently from all the patient and facility variables, 15% of the variation in DSA rate is explained by the center that the patient is treated.

### Sensitivity analysis

Comparing the CPT codes for DSA in the carrier claims files to the DSA procedure codes in the inpatient files of 2016 showed that there were 173 of 246 DSA codes within 14 days of the stroke or the day before. Of these there were 64 not coded as DSA in the inpatient file. So our DSA rate is likely underestimating the true overall DSA rate by approximately 30%. Given that carrier claims were only available for 5% of all claims, though, we were not able to use these data for primary analyses and so the less sensitive ICD procedure codes were used.

## Discussion

This is the first paper to investigate current DSA rate in ischemic stroke patients and explored what patient and facility factors are related to DSA use in these patients. 1.1% of Medicare fee-for-service beneficiaries hospitalized with acute ischemic stroke as primary diagnosis between 2016 and 2020 had a DSA in the 910 hospitals that were able to perform DSA. Against this low overall rate, there is considerable variation between hospitals, with the highest decile of DSA utilizing hospitals performed DSA in more than 2.3%. We did not observe a change in DSA rates over time, even though the MT rate in our study have more than doubled from 2016 to 2020.

It is unclear whether the possible diagnostic benefits of DSA outweigh its risks and cost and, if so, in which ischemic stroke patients this may be the case. Several potential arguments for DSA’s net utility have been advanced. For example, some suggest that whole brain DSA should be performed before or after MT, since it provides additional vessel information compared to CTA, that could change management (finding more than 1 large vessel occlusion, showing a carotid stenosis to be significant and finding asymptomatic aneurysms, that were not diagnosed on pre-intervention CTA).^11^ Others contend that DSA should be performed in every patient that is considered for a carotid intervention, since it would better define whether a stenosis is significant and would actually meet the criteria for a carotid intervention.^12^ Some argue that DSA is needed to screen for rare underlying etiologies such as: Moyamoya, reversible cerebral vasoconstriction syndrome, cerebral vasculitis or dissection.^13^

Our data do not directly measure the rationale of providers for obtaining DSA. Patient and facility factors associated with DSA, likely give some insight into the rationale underlying DSA decisions. In this study we showed that carotid disease (stenosis or occlusion) was one of the strongest factors associated with DSA performance. A carotid procedure was 5 times more likely in patients that had a DSA performed. However, CT-angiography (CTA), MR-angiography (MRA) and ultrasonography, have shown comparable accuracy to DSA, when it comes to carotid stenosis screening.^14–16^ One paper even showed CTA to be superior to DSA, based on histological measurements of removed plaques during carotid endarterectomy.^17^ For intracranial carotid (or more distal) stenosis, CTA has shown to be superior to DSA and MRA.^15^ The other major patient factor associated with DSA was age. We could speculate that arguments to perform DSA in younger patients are a higher prevalence of uncommon causes of stroke and a lower DSA complication risk. And. This is consistent with the observation in these data that uncommon specific stroke causes (dissection, Moyamoya disease, cerebral arteritis, reversible cerebrovascular vasoconstriction syndrome or other cerebrovascular vasospasm and vasoconstriction) were also associated with a higher DSA rate. However, whether DSA is useful in these instances and changes management is unclear, as there is no evidence that DSA outperforms CT-angiography or MR-angiography (including the newer TOF vessel wall imaging techniques).^13^

In addition to the patient factors associated with DSA, specific facility factors were associated with DSA. Most notably, hospitals that perform more MT cases also perform more DSAs. Several possibilities may underlie this finding. It may be the case that that there is more comfort in performing DSA’s in these hospitals and/or through greater procedural experience it may be the case that these facilities have relatively lower procedural risks. Another possibility is that as facilities perform more MTs they come to perceive the angiographic information as important to patient care and thus obtain more diagnostic DSAs. Alternatively, it is possible this may reflect measurement error and that some of these DSAs are actually misclassified MTs or that these DSAs are in some way directly related to MT cases. However, since we excluded DSA’s when dated within 2 days of stroke, we minimized this possibility. We chose this likely conservative cut point to minimize the possibility that DSA procedures were actually a failed attempt for an intervention (MT or carotid artery stent). This choice may have resulted in a small underestimation of DSA rates, but is less likely to bias our evlaution of temporal trends factors associated with DSA. The observation that transfer patients have a higher DSA rate may reflect either unmeasured facility effects — such that hospitals that receive many transfers are more disposed to performing DSA or because they represent a population with an enriched risk of rare stroke etiologies. Another possible factor driving DSA rates at higher utilizing centers are profit motives, as DSAs are generally well remunerated procedures.^18^ This is supported by our observation that-profit hospitals have higher DSA rates Even after considering a wide variety of patient and facility factors, we found considerable variation in the use of DSA at the facility level. 15% of the variance in DSA rate was explained by the specific hospital, after taking all other patient and facility factors into account. Large hospital variation was previously found in DSA use for blunt cervical trauma patients, despite that CTA has already been shown to be the optimal technique with regard to accuracy, safety and cost-effectiveness.^19–21^ So our findings are in line with this study. The hospital variation in our study was mostly explained by the top decile of hospitals of hospitals that perform DSAs more than 4 times as frequently as the median hospital with the capacity to perform DSA.

### Limitations

Given that we used a retrospective database, this leads to inevitable limitations of missing data, and measurement error via miscoding. Our definition of likely diagnostic DSA, to prevent a DSA being actually a failed carotid procedure or MT, may have missed some DSA’s that were done for diagnostic purposes, if they were performed within 2 days after stroke diagnosis. This could have resulted in a lower than actual DSA rate in our study. Also, as observed in our sensitivity analysis, using ICD procedure codes (available in the whole sample) instead of CPT codes (available in 5% of the sample), likely underestimated the DSA rate by about 30%. However, this potential undercounting is less likely to bias our assessment of temporal trends or facility variation. Another potential limitation is that, the Medicare population studied here is not wholly representative of the US health care system. Given the older age distribution of the Medicare population and the observation that age predicts DSA use, our data likely is an underestimate of the overall rate of diagnostic DSA.

#### Final remarks

This study reports recent DSA rates in ischemic stroke patients and patient and facility factors associate with diagnostic DSA. This study does not inform the effect of DSA could on changes in management or outcomes, nor whether DSA’s cost and risk is justified. Prior data has not clearly informed these questions either as there is a lack of data that DSA improves outcome, and guidelines do not state when DSA risk would be acceptable for which specific subgroup of ischemic stroke patients. Considering that DSA has non-negligible risk, high DSA rates can only lead to net benefit if DSA leads to important management changes that improve outcome in a substantial proportion of patients. Therefore DSA must frequently result in markedly beneficial management changes above and beyond what would have happened by only relying on non-invasive imaging. A randomized trial, however, even if it would be performed, may not yield a definitive answer since there is large heterogeneity in patients that undergo DSA to perform DSA and the underlying stroke etiology. Observational data, though, may be able to inform the net benefit question. If we were able to clearly characterize the frequency of DSAs that change evidence-based practices, the net benefit of DSA could be estimated with decision analyses. For now, in the lack of definitive evidence, providers should sparingly and carefully select patients for DSA and consider the elevated complication risks in the, mostly elderly, ischemic stroke population. Additionally, all ischemic stroke patients considered for DSA could be discussed in multi-disciplinary teams (possibly even through the use of multi-disciplinary vascular case conferences), to discuss the possible risks and possible management changes and so help to ensure a reasonable risk-benefit ratio per individual patient.^22^ This is likely particularly important for centers that have a high baseline DSA rate as our data implies that providers at those centers are much more liberal in selecting patients for DSA than at most hospitals that have the capacity to perform DSA.

### Conclusion

DSA is used infrequently in acute ischemic stroke patients and did not change between 2016 to 2020. Hospital variation in DSA use was however large (range: 0.07%-11.1%), and not solely explained by patient and facility factors.

## Data Availability

We used Medicare data that was available for the research group of prof. Burke.

## Acknowledgments

We want to thank Christos Deligkaris for helping Maxim Mulder with helping him access and perform the analysis while visiting Columbus and also after returning to the Netherlands.

